# Major depressive disorder but not bipolar disorder and schizophrenia is a causal factor for type 2 diabetes as determined by Mendelian randomization

**DOI:** 10.1101/2020.09.12.20193060

**Authors:** Heejin Jin, Jeewon Lee, Sohee Oh, Sanghun Lee, Sungho Won

**Affiliations:** Department of Public Health Science, Seoul National University, Seoul, Korea; Medical Research Collaborating Center, Department of Biostatistics, Seoul National University Boramae Hospital, Seoul, Republic of Korea; Department of Psychiatry, Soonchunhyang University Bucheon Hospital, Bucheon, Republic of Korea; Department of Medical Consilience, Graduate School, Dankook University, Yongin-si, Republic of Korea; Institute of Health and Environment, Seoul National University, Seoul, South Korea

## Abstract

**Objective:** In many epidemiologic studies, type 2 diabetes has been reported to be associated with severe mental illness (SMI) such as schizophrenia (SCZ), bipolar disorder (BPD), and major depressive disorder (MDD). However, the relationship between SMI and type 2 diabetes is bi-directional, and the causal relationship remains unclear due to various confounders. Therefore, a Mendelian randomization (MR) study is necessary to identify the causality between them.

**Research Design and Methods:** We conducted a two-sample MR study to identify the causal effect of SMI on type 2 diabetes using the inverse-variance weighted (IVW), MR-Egger, MR-Egger with a simulation extrapolation, weighted median approach, and MR-Pleiotropy RESidual Sum and Outlier methods. The most appropriate method was selected according to the instrument variables assumption.

**Results:** We found that MDD had a significant causal effect on type 2 diabetes from the results obtained using the IVW method (Odds ratio (OR): 1.191, 95% CI: 1.036–1.372, *P* = 0.014); however, this was not observed for BPD (IVW, OR: 1.006, 95% CI: 0.918–1.104, *P* = 0.892) or SCZ (IVW, OR: 1.016, 95% CI: 0.974–1.059, *P* = 0.463). The absence of reverse-causality between MDD and type 2 diabetes was also demonstrated from bi-directional MR studies.

**Conclusions:** These results clearly reveal important knowledge on the causal role of MDD in the risk of type 2 diabetes without a residual confounding, whereas the causality of BPD and SCZ was not shown. Therefore, careful attention should be paid to MDD patients in type 2 diabetes prevention and treatment.

## INTRODUCTION

To date, many epidemiologic studies have suggested the link between type 2 diabetes and severe mental illness (SMI) including bipolar disorder (BPD), major depressive disorder (MDD), and schizophrenia (SCZ) (1). The prevalence of type 2 diabetes among individuals with SMI has been estimated to be 8% to 17% in BPD, and 16% to 25% in SCZ; further, depressed adults have a 37% increased risk of developing type 2 diabetes (2-4). The side effects of the medication used for treating SMI, unhealthy lifestyle behaviors of patients with SMI, and hypothalamic-pituitary-adrenal (HPA) axis dysregulation could contribute to the association of SMI with type 2 diabetes (5; 6). For instance, medications such as antipsychotics, antidepressants, and mood stabilizers are likely to contribute to type 2 diabetes development by leading to insulin resistance or weight gain (1). Moreover, low physical activity, poor diet, smoking, alcohol, and substance abuse in individuals with SMI might lead to type 2 diabetes (7).

In contrast, type 2 diabetes also affects mental health. One meta-analysis indicated that the risk of MDD increases in people with type 2 diabetes (8), and a prospective population-based study showed that the prevalence of schizophrenia was significantly higher in patients with type 2 diabetes than in the general population (9). Further, a cross-sectional study reported that type 2 diabetes and prediabetes may be risk factors in patients with BPD (10). Large longitudinal studies have also demonstrated that type 2 diabetes impacts mental health negatively (11). As the results of epidemiological studies are inconsistent, the true causality between SMI and type 2 diabetes is still unclear with potential biases and confounding factors. Moreover, the difference between the onset age of type 2 diabetes and SMI makes it very challenging to infer a causal relationship. In general, type 2 diabetes affects middle-aged adults (after age 40) with low heritability (10–15%), while SMI usually occurs in young adulthood (in the 20–30s) with high heritability (80–85% for BPD, 80% for SCZ, and 31–42% for MDD) (12; 13). Accordingly, we hypothesized that SMI would precede type 2 diabetes, if there is a causal effect between them.

Therefore, well-designed, two-sample Mendelian randomization (MR) studies, which have widely been adopted using genetic variants such as single nucleotide polymorphisms (SNP) for instrumental variable (IV) analysis are needed to pinpoint the causal relationship between SMI and type 2 diabetes (14). Compared to a one-sample MR, two-sample MR will not lead to inflated type 1 error rated and false-positive findings. To the best of our knowledge, a two-sample MR analysis focusing on SMI and type 2 diabetes is currently lacking. For this purpose, robustness against potential confounders in MR analysis can be achieved if the three core assumptions for IV, including strong association with intermediate exposure, independence with confounders, and no direct path for the outcome are satisfied. We considered several MR approaches including an inverse-variance weighted (IVW) method and sensitivity analyses—MR-Egger, MR-Egger with a simulation extrapolation (SIMEX), weighted median approach, and MR-Pleiotropy RESidual Sum and Outlier (MR-PRESSO) method—in the case of violation of the assumptions (15).

Here, we investigate the causality between SMI and type 2 diabetes using genome-wide summary statistics in a meta-analysis of a large population of European individuals through two-sample MR study.

## RESEARCH DESIGN AND METHODS

### Exposure dataset: summary statistics of genetic association analyses with bipolar disorder

SNPs-BPD associations were obtained from 20,352 cases and 31,358 controls who lived in 14 countries in Europe, North America, and Australia (16). Cases were required to meet international consensus criteria (diagnostic and statistical manual of mental disorders (DSM-IV), internatinal classification of diseases (ICD)-9, or ICD-10) for a lifetime diagnosis of BPD established using structured diagnostic instruments from assessments by trained interviewers, clinician-administered checklists, or medical record reviews. A total of 9,372,253 SNPs were used for genome-wide association (GWA) analyses; among these, 16 genome-wide significant (*P* < 5 x 10^-08^) SNPs associated with BPD were identified after linkage disequilibrium (LD) pruning (distance < 10,000 kb or LD r^2^ < 0.001). After removal of SNPs nominally associated with type 2 diabetes (*P* < 0.05), 11 BPD-associated SNPs were available.

### Exposure dataset: summary statistics of genetic association analyses with major depressive disorder

The summary statistics for association between SNPs-MDD were obtained through large-scale genome-wide association meta-analyses of 807,553 individuals (246,363 cases and 561,190 controls) from the UK Biobank, 23andMe, Inc., and psychiatric genomics consortium (PGC) (17). The definition of MDD was different in each cohort. In the UK Biobank, three MDD phenotypes were used; 1) self-reported help-seeking for problems with nerves, anxiety, tension, or depression, 2) self-reported depressive symptoms, and 3) MDD identified from hospital admission records. In case of 23andMe, Inc., a self-reported clinical diagnosis of depression was used. Finally, cases were required to meet international consensus criteria (DSM-IV, ICD-9, or ICD-10) in PGC. A total of 8,098,588 SNPs were combined for meta-analysis and 50 SNPs were selected as candidates of IVs. None of them were in the same LD block (r^2^ < 0.001) or within 10,000 kb of an established signal. We checked the pleiotropic effect of those SNPs, and eight SNPs associated with type 2 diabetes (*P* < 0.05) were eliminated. Thus, 42 MDD-associated SNPs were used for our analyses.

### Exposure dataset: summary statistics of genetic association analyses with schizophrenia

SNPs-SCZ associations were obtained from 33,640 cases and 43,456 controls from the PGC (18). Cases with clinical diagnosis (not self-report) of SCZ or schizoaffective disorder were included in our study. Summary statistics of GWA analyses for 13,942,226 SNPs were available, and 83 variants that were genome-wide significantly associated with schizophrenia were significant after LD pruning (distance < 10,000 kb or LD r^2^ < 0.001). Eleven SNPs were associated with type 2 diabetes (*P* < 0.05), and the remaining 72 SCZ-associated SNPs were used for our MR analyses.

### Outcome dataset: summary statistics of genetic association analyses with type-2 diabetes

We obtained summary statistics for associations between SNPs-type 2 diabetes from the DIAbetes Genetics Replication And Meta-analysis (DIAGRAM) Consortium stage 1 meta-analyses with 26,676 cases and 132,532 controls (19). Type 2 diabetes diagnosis was based on diagnostic fasting glucose (≥ 7 mmol/L) or HbA1c levels (≥ 6.5%), hospital discharge diagnosis, use of oral diabetes medication, or self-report. Summary statistics of GWA analyses for 12.1 million SNPs were available and were considered for our MR analyses.

### Mendelian randomization analysis

All SNPs associated with BPD, MDD, or SCZ were selected separately as candidates of IVs that had genome-wide significance and were not in a LD block (r^2^ < 0.001) or within 10000 kb of an established signal. *F*-statistics provided an indication of instrument strength and *F>* 10 indicated that the analysis was unlikely to suffer from weak instrument bias (20). A degree of violation of the “NO Measurement Error” (NOME) assumption was quantified using *I*^2^ statistics and *I*^2^ > 90 indicated lesser dilution of the estimates in MR analysis (21). For the detection of pleiotropic outlier SNPs, Cochran’s Q-test in the inverse-variance weighted (IVW) method and Rucker Q’ statistics in the MR-Egger were used (22). We further conducted an MR-Pleiotropy RESidual Sum and Outlier (MR-PRESSO) test as an indicator of no violations of MR assumptions in the final IV sets (23). Given that no weak instrument bias *(F* >10) was observed and the three tests (Cochran’s Q-test, Rucker Q’ test, and MR-PRESSO test) indicated no directional pleiotropic bias, the IVW method was applied, which is robust when all SNPs are valid instruments (15). Here “valid” means that the following three conditions are satisfied: (i) IVs are strongly associated with exposure, (ii) IVs independent of confounders, and (iii) IVs do not affect the outcome directly. If we assume instruments-outcome association and exposure-outcome association are denoted by *β_GY_* and *β_XY_*, respectively, then the IVW estimate of causal effect (i.e., effect of the exposure on the outcome) can be obtained from the inverse-variance weighted mean of ratio estimates(*β_GY_*/*β_XY_*). If the pleiotropy and outlier SNPs were detected from MR-PRESSO, IVW is not recommended, and we consider several sensitivity analyses to minimize bias. Weighted median method provides valid causal estimates unless more than 50% of the instruments are invalid (15). MR-Egger method can estimate appropriate causal effects in the presence of pleiotropy effects even if all SNPs are invalid (15). When Cochran’s Q-test is rejected or both Cochran’s Q and Rucker Q’ tests are rejected, the MR-Egger method is recommended (15). If the measurement error of the instruments is large *(I*^2^ > 90), the method of SIMEX was applied to correct attenuation bias (21). Furthermore, we conducted a bi-directional MR to investigate the presence of reverse-causality between SMI and type 2 diabetes. The Šidák correction for multiple comparisons was used for analysis and thus, threshold of *P* of 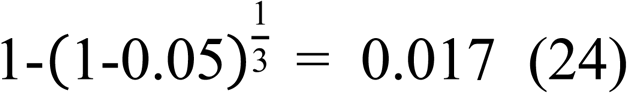 (24). Observed power (or post-hoc power) calculations were performed using the online tool (https://sb452.shinyapps.io/power/) (25). The proportion of variance in the exposure explained by the genetic variants (R^2^) were required for MR power analysis, and 0.23 (BPD), 0.089 (MDD), 0.24 (SCZ), and 0.196 (type 2 diabetes) were used.

## RESULTS

### Effect of bipolar disorder on type 2 diabetes

Eleven SNPs that genome-wide significantly associated with BPD but not type 2 diabetes were used as instrument variables. All SNP-exposure and SNP-outcome effects are presented in Supplementary Table 1. We found no evidence of weak instrument bias (*F*-statistic = 32.9), and heterogeneity and outlier pleiotropy were not observed (Q-test, *P* = 0.718; Q’-test, *P* = 0.821; MR-PRESSO global test, *P* = 0.695) (Table 1). The MR-Egger test also indicated no directional pleiotropic bias (intercept *P* = 0.165) and there was no violation of the NOME assumption (*I*^2^ = 96.9%) (Table 1). Since all IV assumptions were satisfied, the IVW method was considered to be the most appropriate to provide unbiased estimate (15; 22). IVW showed non-significant effect on the type 2 diabetes (odds ratio (OR): 1.006, 95% CI: 0.918–1.104, *P =* 0.892). The post-hoc statistical power estimated using OR was 2.5%. Non-significant results were also obtained from other sensitivity analyses (MR-Egger, OR: 0.681, 95% CI: 0.389–1.191, *P =* 0.178; MR-Egger (SIMEX), OR: 0.995, 95% CI: 0.908–1.089, *P =* 0.917; weighted median, OR: 0.982, 95% CI: 0.868–1.111, *P* = 0.770) (Table 2). We checked the reverse-causal association between BPD and type 2 diabetes with the 35 SNPs associated with type 2 diabetes. There was no weak instrument bias (*F*-statistic = 35.8) and no violation of the NOME assumption (*I^2^* = 97.3%). No evidence of heterogeneity was found from the Q-test (*P* = 0.684), Q’-test (*P* = 0.639), and MR-PRESSO global test (*P* = 0.598) (Table 1). Non-significant reverse-causal effect was found (IVW, OR: 1.031, 95% CI: 0.971–1.095, *P* = 0.313; MR-Egger, OR: 1.051, 95% CI: 0.793–1.394, *P*=0.727; MR-Egger (SIMEX), OR: 0.993, 95% CI: 0.909–1.083, *P =* 0.867; weighted median OR: 1.041, 95% CI: 0.956–1.134, *P* = 0.349) (Table 3). The associations of the variants with BPD and type 2 diabetes are shown in a scatter plot with four MR-fitted lines (Figure 1A).

**Table 1.** Assumption test for instrumental variable sets

| No. | Exposure | Outcome | N | F-stat | *I*<sup>2</sup> (%) | Q-test | Q'-test | MR-PRESSO<br>global test |
| --- | --- | --- | --- | --- | --- | --- | --- | --- |
| 1 | BPD | type 2<br>diabetes | 11 | 32.9 | 96.9 | 0.718 | 0.821 | 0.695 |
| 2 | MDD |  | 42 | 37.9 | 97.3 | 0.808 | 0.788 | 0.733 |
| 3 | SCZ |  | 72 | 41.5 | 97.6 | 0.319 | 0.290 | 0.704 |
| 4 | type 2<br>diabetes | BPD | 35 | 35.8 | 97.3 | 0.684 | 0.639 | 0.598 |
| 5 |  | MDD | 39 | 36.0 | 97.3 | 0.714 | 0.683 | 0.711 |
| 6 |  | SCZ | 43 | 35.8 | 97.2 | <0.001 | <0.001 | <0.001 |

N = Number of instruments, F-stat = F statistics, Q-test = *P*-value for the Q-test from IVW, Q'-test = *P*-value for the Q'-test from MR-Egger, MR-PRESSO global test = *P*-value for MR-PRESSO global test, BPD = bipolar disorder, MDD = major depressive disorder, SCZ = schizophrenia, MR- PRESSO = MR-Pleiotropy RESidual Sum and Outlier

**Figure 1.**
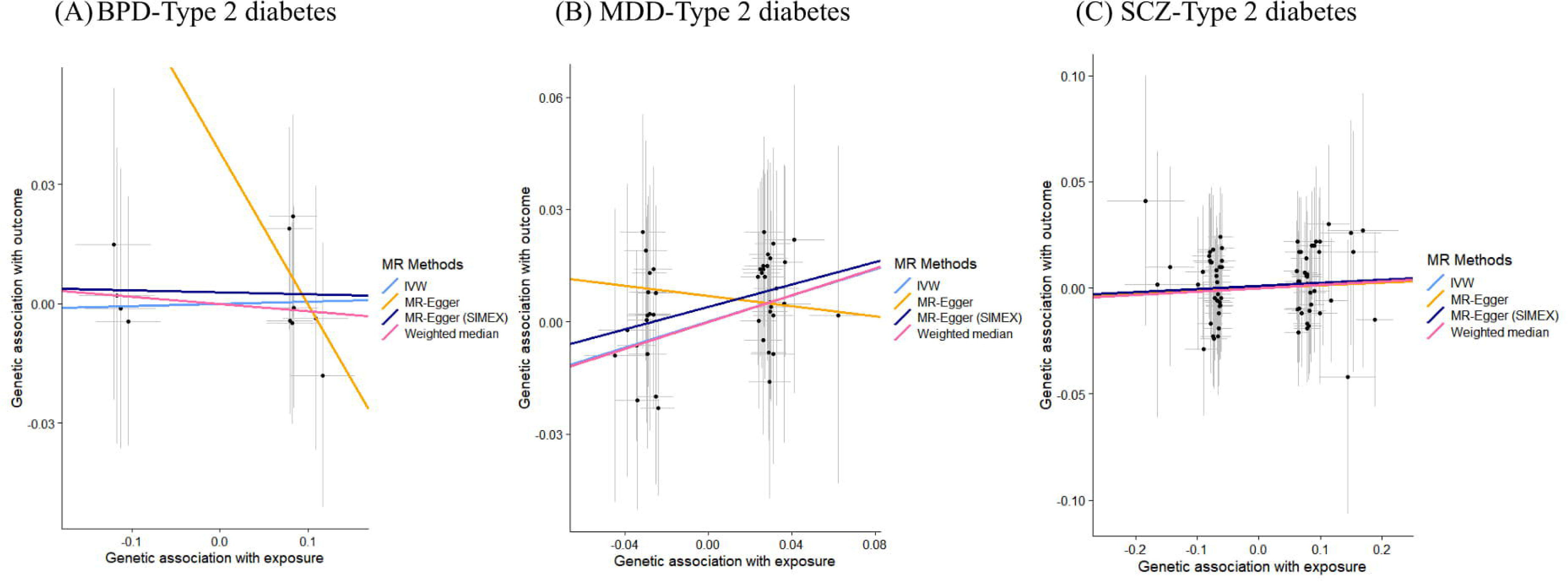
Scatter plots showing the associations of the genetic variants with psychiatric disorders and type 2 diabetes for different methods of Mendelian randomization. (A) The associations of the variants with BPD and type 2 diabetes. (B) The associations of the variants with MDD and type 2 diabetes. (C) The associations of the variants with SCZ and type 2 diabetes. Abbreviations: BPD = bipolar disorder, MDD = major depressive disorder, SCZ = schizophrenia, MR = Mendelian randomization, IVW = inverse-variance weighted, SIMEX = simulation extrapolation

**Table 2.** Mendelian randomization results

| MR methods | parameter | N | OR | 95% CI | P | Power |
| --- | --- | --- | --- | --- | --- | --- |
| 1. Effect of BPD on type 2 diabetes |  |  |  |  |  |  |
| IVW | Estimate | 11 | 1.006 | [0.918, 1.104] | 0.892 | 2.5% |
| MR-Egger | Intercept |  | 1.039 | [0.985, 1.095] | 0.165 | 100% |
|  | Slope |  | 0.681 | [0.389, 1.191] | 0.178 | - |
| MR-Egger (SIMEX) | Intercept |  | 1.003 | [1.010, 1.994] | 0.540 | 2.1% |
|  | Slope |  | 0.995 | [0.908, 1.089] | 0.917 | - |
| Weighted median | Estimate |  | 0.982 | [0.868, 1.111] | 0.770 | 13.8% |
| MR-PRESSO | Estimate |  | No outlier |  | - | - |
| 2. Effect of MDD on type 2 diabetes |  |  |  |  |  |  |
| IVW | Estimate | 42 | 1.191 | [1.036, 1.372] | 0.014 | 100% |
| MR-Egger | Intercept |  | 1.007 | [0.982, 1.033] | 0.566 | 74% |
|  | Slope |  | 0.934 | [0.402, 2.172] | 0.875 | - |
| MR-Egger (SIMEX) | Intercept |  | 1.004 | [1.000, 1.008] | 0.036 | 100% |
|  | Slope |  | 1.162 | [1.024, 1.318] | 0.025 | - |
| Weighted median | Estimate |  | 1.196 | [0.984, 1.453] | 0.072 | 100% |
| MR-PRESSO | Estimate |  | No outlier |  | - | - |
| 3. Effect of SCZ on type 2 diabetes |  |  |  |  |  |  |
| IVW | Estimate | 72 | 1.016 | [0.974, 1.059] | 0.463 | 10.9% |
| MR-Egger | Intercept |  | 1.000 | [0.987, 1.014] | 0.969 | 6.5% |
|  | Slope |  | 1.012 | [0.851, 1.204] | 0.888 | - |
| MR-Egger (SIMEX) | Intercept |  | 1.001 | [0.998, 1.004] | 0.427 | 9.6% |
|  | Slope |  | 1.015 | [0.981, 1.060] | 0.511 | - |
| Weighted median | Estimate |  | 1.016 | [0.955, 1.081] | 0.615 | 10.9% |
| MR-PRESSO | Estimate |  | No outlier |  | - | - |

N = Number of instruments, BPD = bipolar disorder, MDD = major depressive disorder, SCZ = schizophrenia, MR = Mendelian randomization, IVW = inverse-variance weighted, MR- PRESSO = MR-Pleiotropy RESidual Sum and Outlier, SIMEX = simulation extrapolation, OR = odds ratio

**Table 3.** Bi-directional Mendelian randomization results

| MR methods | parameter | N | OR | 95% CI | P | Power |
| --- | --- | --- | --- | --- | --- | --- |
| 1. Effect of type 2 diabetes on BPD |  |  |  |  |  |  |
| IVW | Estimate | 35 | 1.031 | [0.971, 1.095] | 0.313 | 18.9% |
| MR-Egger | Intercept |  | 0.998 | [0.974, 1.022] | 0.890 | 52.7% |
|  | Slope |  | 1.051 | [0.793, 1.394] | 0.727 | - |
| MR-Egger (SIMEX) | Intercept |  | 0.997 | [0.989, 1.004] | 0.428 | 2.1% |
|  | Slope |  | 0.993 | [0.909, 1.083] | 0.867 | - |
| Weighted median | Estimate |  | 1.041 | [0.956, 1.134] | 0.349 | 34.3% |
| MR-PRESSO | Estimate |  | No outlier |  | - | - |
| 2. Effect of type 2 diabetes on MDD |  |  |  |  |  |  |
| IVW | Estimate | 39 | 1.002 | [0.985, 1.020] | 0.793 | 2.2% |
| MR-Egger | Intercept |  | 0.999 | [0.994, 1.005] | 0.852 | 18.6% |
|  | Slope |  | 1.008 | [0.949, 1.070] | 0.798 | - |
| MR-Egger (SIMEX) | Intercept |  | 1.001 | [0.999, 1.002] | 0.278 | 2.2% |
|  | Slope |  | 1.002 | [0.985, 1.019] | 0.808 | - |
| Weighted median | Estimate |  | 1.007 | [0.982, 1.033] | 0.581 | 14% |
| MR-PRESSO | Estimate |  | No outlier |  | - | - |
| 3. Effect of type 2 diabetes on SCZ |  |  |  |  |  |  |
| IVW | Estimate | 43 | 1.006 | [0.939, 1.075] | 0.872 | 2.2% |
| MR-Egger | Intercept |  | 0.997 | [0.974, 1.019] | 0.783 | 50.1% |
|  | Slope |  | 1.040 | [0.806, 1.343] | 0.758 | - |
| MR-Egger (SIMEX) | Intercept |  | 1.003 | [0.997, 1.009] | 0.246 | 3.3% |
|  | Slope |  | 1.009 | [0.941, 1.082] | 0.785 | - |
| Weighted median | Estimate |  | 0.980 | [0.914, 1.050] | 0.564 | 12.4% |
| MR-PRESSO | Estimate | 41 | 0.999 | [0.991, 1.009] | 0.987 | 1% |

N = Number of instruments, BPD = bipolar disorder, MDD = major depressive disorder, SCZ = schizophrenia, MR = Mendelian randomization, IVW = inverse-variance weighted, MR- PRESSO = MR-Pleiotropy RESidual Sum and Outlier, SIMEX = simulation extrapolation, OR = odds ratio

### Effect of major depressive disorder on type 2 diabetes

Forty-two independent SNPs that associated with MDD but not type 2 diabetes were used as IVs. All SNP-exposure and SNP-outcome effects are presented in Supplementary Table 2. Weak instrument assumption was guaranteed by *F*-statistic (37.9) and there was no measurement error of estimates from the MR study (*I^2^* = 97.3%). No evidence of heterogeneity or pleiotropy was confirmed through the Q-test (*P* = 0.808), Q’-test (*P* = 0.788), and MR-PRESSO global test (*P* = 0.733) (Table 1). Since all IV assumptions were satisfied, the IVW method was chosen and showed significant results (OR: 1.191, 95% CI: 1.036–1.372, *P* = 0.014). The post-hoc statistical power estimated from the ORs was 100%. Among the other sensitivity analyses, MR-Egger (SIMEX) showed a consistent significant result (OR: 1.162, 95% CI: 1.024–1.318, *P* = 0.025) and weighted median showed a nominally significant (*P* < 0.1) result (OR: 1.196, 95% CI: 0.984–1.453, *P* = 0.072). Non-significant result of MR-Egger may be attributable to the low power of ME-Egger (OR: 0.934, 95% CI: 0.402–2.172, *P* = 0.875) (Table 2). To check the reverse-causal effect, 39 variants were used as instruments. We found that all assumptions for MR analyses were preserved (*F*-statistic = 36.0; Q-test, *P* = 0.714; Q’-test, *P* = 0.683; MR-PRESSO global test, *P* = 0.711; *I^2^* = 97.3%) (Table 1). Non-significant reverse-causal effect was found (IVW, OR: 1.002, 95% CI: 0.985–1.020, *P* = 0.793; MR-Egger, OR: 1.008, 95% CI: 0.949–1.070, *P =* 0.798; MR-Egger (SIMEX), OR: 1.002, 95% CI: 0.985–1.019, *P =* 0.808; weighted median, OR: 1.007, 95% CI: 0.982–1.134, *P* = 0.581) (Table 3). The associations of the variants with MDD and type 2 diabetes are shown in a scatter plot with four MR-fitted lines (Figure 1B).

### Effect of schizophrenia on type 2 diabetes

Seventy-two independent SNPs that associated with SCZ, but not type 2 diabetes were found to have strong instrument strength (*F*-statistic = 41.5) and non-significant heterogeneity and outlier pleiotropy (Q-test, *P* = 0.319; Q’-test, *P* = 0.290; MR-PRESSO global test, *P* = 0.704) (Table 1). All SNP-exposure and SNP-outcome effects are presented in Supplementary Table 3. In addition, there was no dilution bias from violation of the NOME assumption (*I^2^* = 97.2%). The IVW method was the most powerful and showed a non-significant result (OR: 1.016, 95% CI: 0.974–1.059, *P* = 0.463). The post-hoc statistical power by estimated ORs was 10.9%. The other sensitivity analyses results showed a non-significant effect of SCZ on type 2 diabetes (MR-Egger, OR: 1.012, 95% CI: 0.851–1.204, *P =* 0.888; MR-Egger (SIMEX), OR: 1.015, 95% CI: 0.981–1.060, *P =* 0.511; weighted median, OR: 1.016, 95% CI: 0.955–1.081, *P* = 0.615) (Table 2). To detect reserve-causal effect, forty-three SNPs were used as instruments. *F*-statistic showed no weak instrument bias (*F*-statistic = 35.8) and no violation of the NOME assumption (*I^2^* = 97.2) (Table 1). Heterogeneity test showed substantial evidence of outlier pleiotropy using the Q-test (*P* < 0.001), Q’-test (*P* < 0.001), and MR-PRESSO global test (*P* < 0.001) (Table 1). Since all three tests were rejected, the MR-PRESSO method was adopted (15) after excluding the two outlier SNPs (OR: 0.999, 95% CI: 0.991–1.009, *P* = 0.987), suggesting non-significant effect of type 2 diabetes on SCZ. The associations of the variants with SCZ and type 2 diabetes are shown in a scatter plot with four MR-fitted lines (Figure 1C).

## CONCLUSIONS

In the present study, two-sample MR results provided solid evidence in support of the hypothesis that MDD increases the risk of type 2 diabetes, whereas BPD and SCZ were not identified as risk factors for type 2 diabetes. Bi-directional MR study also confirmed that there was no reverse-causality between SMI and type 2 diabetes, and the asymmetry in the effects of the instruments on type 2 diabetes and MDD supports the fact that MDD is one of the causal factors that influences type 2 diabetes. However, causal effects of MDD on type 2 diabetes development have been demonstrated to be controversial in observational studies. One systematic review demonstrated that MDD is associated with a 60% increased risk of type 2 diabetes, while the evidence is also compatible with the high prevalence rates of MDD among individuals with type 2 diabetes (27). A large meta-analysis showed that type 2 diabetes is associated with only a modestly increased risk of MDD (28). MDD is difficult to detect in older adults which may partially explain why this association was so modest (29).

Our finding, the causal role of MDD in increased risk of type 2 diabetes, could be explained by the pathophysiological mechanisms underlying the two diseases. Two major molecular mechanisms have been suggested to explain the causal pathway between them. First, the HPA axis, a central stress response system is commonly activated in MDD patients suffering from emotional stressors, leading to a rise in the levels of glucocorticoids, primarily cortisols (30). High cortisol level induces and aggravates insulin resistance in a vicious cycle, such as increased β-cell function and increased insulin release to glucose challenge by exacerbating progression to insulin resistance (31). Second, the sympathetic nervous system (SNS) activity is also elevated in MDD (32). The SNS axis interacts complexly with the HPA axis to maintain homeostasis during stress, resulting in an increased release of cortisol and other glucocorticoids, catecholamines, growth hormone, and glucagon. Indeed, the catecholamines have marked metabolic effects, particularly on glucose metabolism (33).

However, our findings are inconsistent with some observational study suggesting a causal role of BPD and SCZ in the risk of type 2 diabetes and that type 2 diabetes predicts the development of MDD (3). Such associations may have been driven by residual confounding, because many aspects of the relationship between SMI and type 2 diabetes are yet to be examined in a controlled manner, and there are many suggestive evidences that can act as confounders. First, sedentary lifestyle and low physical activity, which have been demonstrated to be strongly associated with SMI, may play a role as a potential confounder (1). A large meta-analysis of general population studies reported that sedentary behavior is independently associated with an increased risk of type 2 diabetes (34). In addition, side effects of medication could also be another important potential confounder. A systematic review of cross-sectional and prospective studies of psychotropic medications and physical diseases indicated that the use of antipsychotics, antidepressants, and mood stabilizers can contribute to an increased BMI which is a major risk factor for type 2 diabetes (35). Especially, antipsychotics, such as clozapine and olanzapine; antidepressants, such as paroxetine; and mood stabilizers, such as lithium and valproate have been associated with increasing obesity. Further, the prevalence of smoking behaviors (daily smoking) was the highest in SCZ, followed by BPD and MDD compared with the general population and the association with smoking is very strong in SCZ and BPD, while it is less strong in MDD (36). The evidence that nicotine addiction begins before any of these SMIs develop suggests that there are shared genes associated with nicotine addiction and SMI (37).

MR studies on the association between SMI and type 2 diabetes are scarce, with no studies on BPD with type 2 diabetes. To investigate the potential causal relationship of type 2 diabetes with MDD, MR analysis was performed with a large longitudinal cohort from 2011 to 2013 in China (38). Genetic risk scores for type 2 diabetes were chosen as the instruments and two-stage multiple regression was used for statistical analysis. The results provided evidence of a potential causal effect of type 2 diabetes on MDD, which is the opposite of our results. We thought that there is a finite-sample bias in the existing research, because in a one-sample setting, the fitted values from the first-stage regression are correlated with the outcome in finite samples even in the absence of a causal effect (39). Regarding the MR studies of SCZ and type 2 diabetes, two-sample MR was performed using the IVW and MR-Egger methods in Europeans, East Asians, and trans-ancestry groups (40). No evidence of a causal role of type 2 diabetes for SCZ was observed in any of the analyses, which is consistent with our findings; however, they did not perform bi-directional analysis for causal effect of SCZ on type 2 diabetes. Unlike epidemiological studies, the previous and the present MR studies could not consider multi-episode status of disease, which may have led to non-significant results of SCZ and BPD. This could be because multi-episode (versus first-episode) persons with SMI were significantly more likely to have type 2 diabetes than matched controls in the meta-analysis of observational studies (1).

Several limitations of our study need to be acknowledged. First, there are different clinical subtypes of MDD (melancholic, psychotic, atypical, or undifferentiated) and bipolar disorder (type 1 or 2) and mood states (manic, depressive, mixed, or euthymic); however, a large category of diseases was analyzed without distinction. Second, although we conducted bi-directional MR studies, the sample size of GWA studies for instruments and BPD/SCZ (less than 100,000) was relatively small, which could lead to low power of the analysis. Third, we only included European population; hence, it is difficult to apply the same clinical interpretation to other races. Nevertheless, the present study has implications in that the well-designed, two-sample MR study for SMI and type 2 diabetes produced less biased results than those of the existing epidemiological studies or one-sample MR study. In addition, the present study clearly showed the causality of MDD on type 2 diabetes, which is supported by previously reported underlying biological mechanisms. Therefore, it is imperative to consider screening for diabetes and metabolic abnormalities in patients with MDD or probable MDD.

## Data Availability

The data can be downloaded from the following link.

https://www.med.unc.edu/pgc/download-results/

http://diagram-consortium.org/downloads.html

## ACKNOWLEDGEMENTS

This work was supported by the Industrial Core Technology Development Program (20000134) funded by the Ministry of Trade, Industry and Energy (MOTIE, Korea), and the National Research Foundation of Korea (2017M3A9F3046543). The authors declare that the research was conducted in the absence of any commercial or financial relationships that could be construed as a potential conflict of interest.

## AUTHOR CONTRIBUTIONS

H.J. analyzed and interpreted the results and wrote the manuscript. J.L and S.O contributed to discussion. S.W. and S.L. designed the study. All authors revised this paper critically for important intellectual content.

